# Nested PCR for the detection of *Taenia solium* DNA in stool samples

**DOI:** 10.1101/2020.06.29.20142950

**Authors:** C Franco-Muñoz, A Arévalo, S Duque-Beltran

## Abstract

The traditional parasitological method to diagnose taeniasis is the microscopic observation of eggs in stool samples. However, this method does not allow differentiation between *Taenia saginata* and *Taenia solium*. This aim of this study was to achieve the detection of *T. solium* DNA by polymerase chain reaction (PCR) and to evaluate the cross-reaction with other species of the genus *Taenia* and other intestinal parasites. DNA was extracted from adult *T. solium* cestodes by cryolysis in liquid nitrogen and with the DNA stool extraction kit from Qiagen. The detection limit of the test was evaluated by DNA dilutions in water and in stool samples. DNA was extracted from proglottids of *T. saginata* and *T. crassiceps* and from stool samples containing other intestinal parasites using ethanol treatment, alkaline lysis, and the DNA stool extraction kit. Nested PCR was used to amplify a previously described fragment of the *Tso31* gene, and the PCR products were analyzed by electrophoresis in 2% agarose gels followed by staining with GelRed. The nested PCR of the *Tso31* gene allowed the detection of *T. solium* DNA in stool samples with a detection limit of 20 pg of parasite DNA. PCR showed no cross-reaction with *T. saginata, T. crassiceps*, or other intestinal parasites of public health importance in Colombia.

## INTRODUCTION

Diseases of the taeniasis/cysticercosis complex are a public health problem in countries with ecoepidemiological conditions conducive to the maintenance and transmission of these parasites, such as Colombia (1). Taeniasis/cysticercosis is associated with poverty and health illiteracy in communities (2). Taeniasis is caused by the ingestion of larvae (cysticerci), and two species have been identified to infect humans: *Taenia saginata* (beef tapeworm) and *Taenia solium* (pork tapeworm). Ingestion of *T. solium* eggs is the cause of neurocysticercosis, a major human health problem, especially due to its main complication, epilepsy (3). *T. solium* infections cause approximately 28,000 deaths worldwide each year, and approximately 2.7 million years of life are lost due to disability (4).

Identifying people infected with *T. solium* is important to administer timely treatment and interrupt the transmission cycle of the parasite. The traditional diagnostic method is the direct observation of parasite eggs by conventional microscopy in stool samples. However, due to the biological characteristics of the release of *T. solium* eggs in feces, the sensitivity of the microscopic method can vary between 3.9% (5) and 52.5% (6). Microscopy does not differentiate between *T. saginata* and *T. solium* since the eggs are morphologically similar.

Diagnosis based on the detection of *T. solium* DNA by polymerase chain reaction (PCR) is a very useful technique with 100% specificity and 97-100% sensitivity when a fragment of the *Tso31* gene of *T. solium* is amplified by nested PCR (7, 8). The objective of this study was to achieve the detection of *T. solium* DNA by PCR in stool samples and to evaluate the cross-reaction with other species of the genus *Taenia* and other intestinal parasites.

## METHODS

### Ethical aspects

This study was approved and monitored by the Ethics Committee of National Health Institute of Colombia in accordance with national regulations and the Declaration of Helsinki. The patients signed an informed consent statement for the use of the stool sample. The animal experiments were performed following the regulations and ethical codes of the Barrier Bioterium and Experimentation BSL2 of the National Institute of Health of Colombia (Declaration of Helsinki) and according to the three principles of laboratory animal experimentation (reduce, refine, replace).

### Biological material

#### In vivo maintenance of T. solium in an animal model

Golden hamsters were infected with viable *T. solium* larvae (cysticerci) as described (9). The animals were infected with five cysticerci orally and then subjected to immunosuppression with methylprednisolone (2 mg/animal) for 15 days. After four weeks, the hamsters were sacrificed in a CO_2_ chamber, and the adult forms of *T. solium* were recovered from the intestines, which were lyophilized and stored at −80 °C. All animals were female.

#### Stool sample from uninfected hamsters

Uninfected hamsters were sacrificed in a CO_2_ chamber, and stool sample was recovered from the large intestine. The samples were stored in absolute ethanol at room temperature.

#### Parasitologically cheked stool samples with intestinal parasites

Seven samples of fecal matter preserved in formalin-glycerol stored in the Biological Bank of the Parasitology Group of the National Institute of Health of Colombia were used. The samples were parasitologically cheked proven to have the following parasites by conventional microscopy: eggs of *Taenia* spp., eggs of *Hymenolepis diminuta*, eggs of *Hymenolepis nana*,eggs of *Ascaris lumbricoides*, eggs of *Trichuris trichiura*, eggs of *Uncinaria*, cysts of *Giardia*, cysts of *Chilomastix mesnili*, cysts of *EntamoebaHartmanni*, cysts of *Entamoeba coli*,, cysts of *Endolimax. nana*, cysts of *Entamoebahistolytica/Entamoeba dispar* complex and *Blastocystis hominis*.

#### Biological material for cross-reaction tests

Biological material from related parasites isolated and stored in the Biological Bank of the Parasitology Group of the National Institute of Health of Colombia was used. Samples of adult-stage *T. crassiceps* and *Fasciola hepatica* lyophilized and stored at −80 °C as well as proglottids of *T. saginata* and eggs of *Strongyloides stercoralis* preserved in 5% formalin-glycerol at room temperature were included.

### DNA extraction

#### Obtaining DNA from lyophilized parasites

Cryolysis was performed in liquid nitrogen in a porcelain flask, and the DNA was extracted with the QIAamp DNA stool mini kit from Qiagen following the manufacturer’s instructions. The quantity and quality of the extracted DNA were evaluated with a Nanodrop 2000 spectrophotometer (Thermo Scientific).

#### Obtaining DNA from samples preserved in formalin

The protocol described by Hykin et al. (2015) was followed (10). Five hundred microliters of each sample was centrifuged at 16,000 × g for 5 minutes to remove formalin. Next, treatment was performed with decreasing concentrations of ethanol (100% to 70%) to hydrate the DNA. The samples were treated with 0.5 M EDTA for 1 hour at 55 °C to inhibit DNases and then lysed with ASL buffer (Qiagen) and proteinase K for 2 hours at 55 °C. After this step, the extraction protocol of the QIAamp DNA stool mini kit was continued following the manufacturer’s instructions (Qiagen). The quantity and quality of the extracted DNA was evaluated with a Nanodrop 2000 spectrophotometer (Thermo Scientific).

#### Obtaining DNA from parasites preserved in formalin-glycerol

Biological material (proglottids or eggs) was centrifuged at 16,000 × g for 5 minutes to remove formalin. After that, treatment was performed with decreasing concentrations of ethanol (100% to 70%) to hydrate the DNA. DNA was extracted according to a high-temperature alkaline lysis protocol described by Campos et al. (2012) (11) using 0.5-mm glass beads during lysis, followed by purification using the QIAamp DNA stool mini kit according to the manufacturer’s instructions.

#### Obtaining DNA from stool samples preserved in ethanol

A total of 200 µl of fecal matter preserved in formalin was collected and centrifuged at 16,000 × g for 5 minutes. Then, 1.4 ml of ASL lysis buffer (Qiagen) was added to the precipitate, and DNA extraction was continued using the QIAamp DNA stool mini kit following the manufacturer’s instructions.

### Nested PCR of the Tso31 gene

The first PCR was performed based on the method described by Mayta et al. in 2008 (8) with the following primers that yielded a 691-bp amplified fragment of the *Tso31* gene of *T. solium*:

*Forward primer F1* 5’-ATGACGGCGGTGCGGAATTCTG-3’

*Reverse* p*rimer R1* 5’-TCGTGTATTTGTCGTGCGGGTCTA-3’

PCR was performed with 20 ng of DNA, 1X amplification buffer (20 mM Tris-HCl, 50 mM KCl), 3 mM MgCl_2_, 200 µM of each dNTP, 0.8 µM of each primer, and 0.125 U of Platinum *Taq* DNA polymerase (Tucan *Taq*) in a final volume of 20 µl.

The product of the first round of PCR was used as a template to perform the nested PCR as described by Mayta et al. (2008) (8). The following primers were used to obtain an amplified 234-bp fragment:

*Forward primer F589* 5’-GGTGTCCAACTCATTATACGCTGTG-3’

*Reverse primer* R294 5’-GCACTAATGCTAGGCGTCCAGAG-3’

The nested PCR was performed with 1 µl of the first PCR amplification product, 1X amplification buffer (20 mM Tris-HCl, 50 mM KCl), 2.5 mM MgCl_2_, 200 µM of each dNTP, 0.8 µM of each primer, and 0.125 U of platinum *Taq* polymerase (Tucan *Taq*) in a final volume of 20 µl.

A C1000 thermal cycler (Bio-Rad) was used for PCR with the following thermal profile. First PCR: 95 °C for 3 minutes followed by 25 amplification cycles of 95 °C for 30 seconds, 55 °C for 30 seconds, and 72 °C for 1 minute. Second PCR: 95 °C for 3 minutes followed by 40 amplification cycles of 95 °C for 30 seconds, 50 °C for 30 seconds, and 72 °C for 1 minute.

The PCR products were subjected to agarose gel electrophoresis to visualize the amplified fragments. Five microliters of the amplified product was loaded onto a 2% agarose gel in TBE buffer solution (Tris, boric acid, EDTA) and 4 µl of GelRED. Electrophoresis was performed at 80 V for 1 hour, and the gel was visualized using the Gel Doc imager and Image Lab software (Bio-Rad).

### Limit of detection of nested PCR

Ten-fold dilutions were performed with *T. solium* DNA starting from a concentration of 20 ng/µl and going to 20 fg/µl. The diluted DNA was used as a template to perform the nested PCR of the *Tso31* gene as described above.

### Performance of the PCR in samples

A total of 200 µl of uninfected hamster fecal matter preserved in absolute ethanol was mixed with *T. solium* DNA in different amounts between 2 µg and 20 pg. Then total DNA extraction was performed as described above, and the *Tso31* gene of *T. solium* was amplified to evaluate the limit of detection of the PCR in a sample matrix similar to that expected for clinical samples.

## RESULTS

It was possible to reproduce the PCR conditions reported by Mayta et al. (2008) using as template the DNA extracted from the adult form of *T. solium* recovered from intestines of infected hamsters in the laboratory. The amplified fragments of the *Tso31* gene in the first and second PCR rounds matched the expected size as reported in the literature and predicted by *in silico* analysis (Fig. 1A). Nested PCR can specifically amplify the DNA of *T. solium* without cross-reacting with other related species of the genus *Taenia*, such as *T. saginata* and *T. crassiceps* (Fig. 1B), or with other intestinal parasites, such as *Strongyloides stercoralis* and *Fasciola hepatica* (Fig. 1C).

**Fig. 1.**
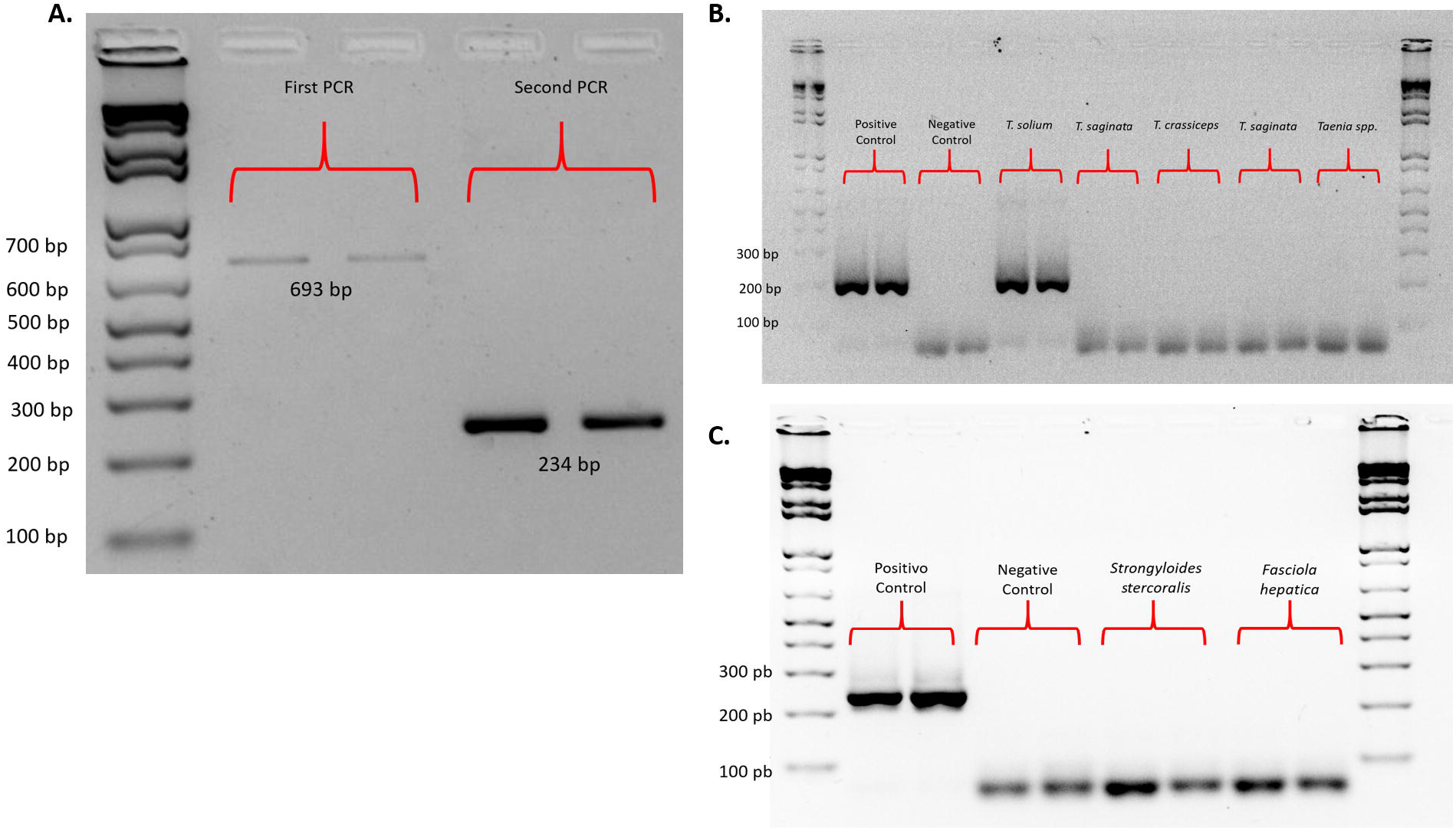
Nested PCR specifically amplifies the DNA of *T. solium* and not that of *T. saginata, T. crassiceps*, or other intestinal parasites. **A**. Amplification of *Tso31* gene fragments using the conditions described by Mayta et al. and *T. solium* DNA as template. **B**. Amplification of the *Tso31* gene by nested PCR using DNA from different species of the genus *Taenia* recovered from humans and mice as templates. **C**. Amplification of the *Tso31* gene by nested PCR using DNA from *Strongyloides stercoralis* and *Fasciola hepatica* isolated from humans. All experiments were performed in duplicate. Positive control: DNA extracted from *T. solium* in adult form recovered from hamster. Negative control: reaction mixture without template.

The limit of detection of PCR in stool samples containing *T. solium* DNA was a proportion of 20 ng of DNA in 200 µl of stool sample (Fig. 2A). However, considering that the DNA was added before extraction, the PCR performance may have been influenced by the presence of DNases in the samples, PCR inhibitors, and the extraction yield. To evaluate the PCR performance under the best possible conditions, serial dilutions of *T. solium* DNA were performed in water, and amplification of the *Tso31* gene fragment was recorded with up to 20 pg of DNA (Fig. 2B), suggesting that the PCR limit of detection varied between 20 ng and 20 pg of DNA, depending on the sample characteristics.

**Fig. 2.**
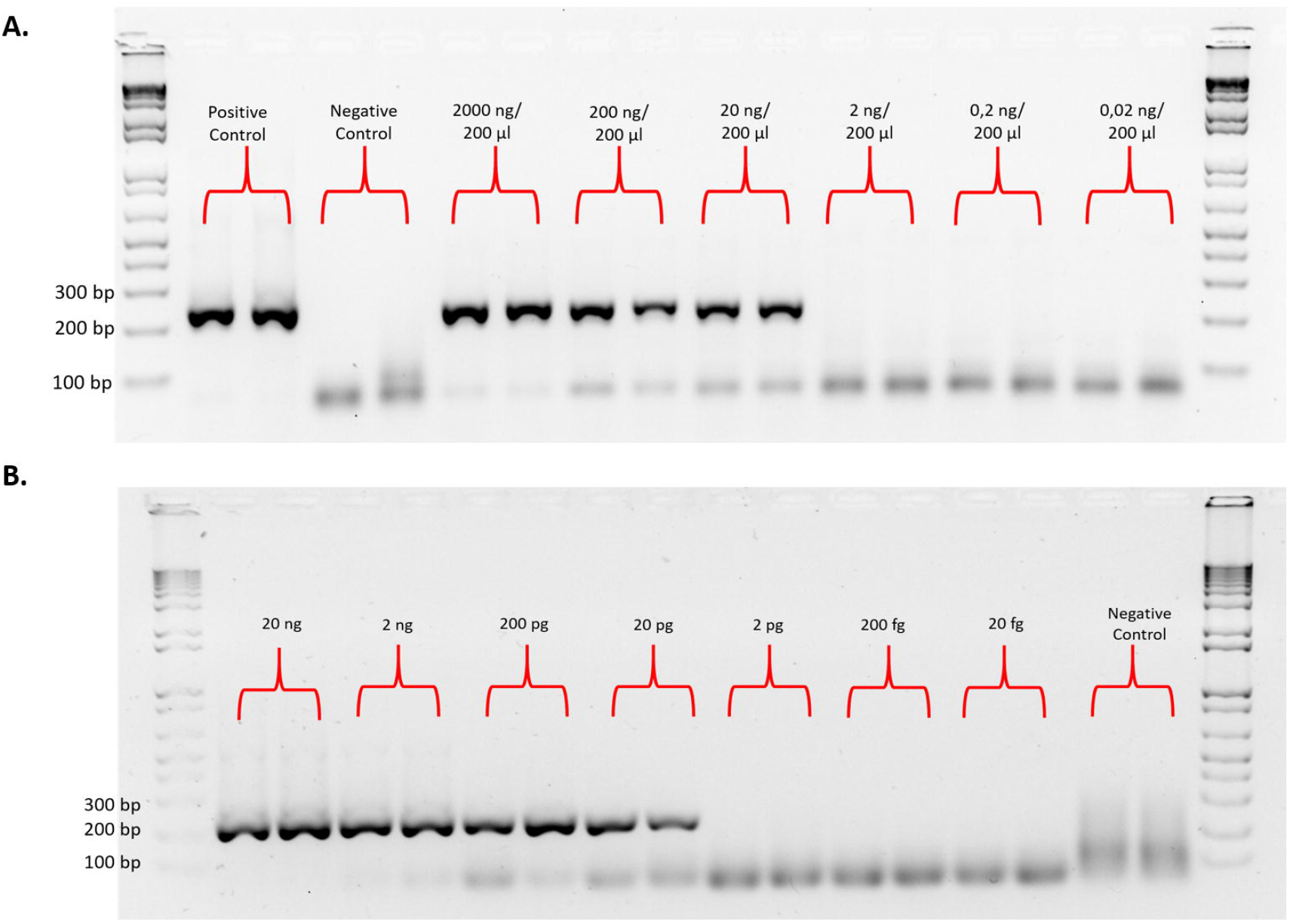
*Tso31* nested PCR has a detection limit of 20 pg of DNA in water and 20 ng in 200 µl of fecal matter. **A**. Amplification of a fragment of the *Tso31* gene by nested PCR in samples of uninfected hamster fecal matter preserved in absolute ethanol with *T. solium* DNA in different amounts between 2 µg and 20 pg. All experiments were performed in duplicate. Positive control: DNA extracted from *T. solium* in adult form recovered from hamster. Negative control: reaction mixture without template. **B**. Amplification of a fragment of the *Tso31* gene by nested PCR in different dilutions of *T. solium* DNA in water between 20 ng and 20 fg.

The results of the nested PCR of the *Tso31* gene in DNA extracted from human stool samples with microscopic verification of *Taenia* spp. and other intestinal parasites are shown in Fig. 3. All samples were negative, suggesting that the nested PCR test did not cross-react with any of the parasites present in the different samples (Fig. 3A). In the clinical samples with microscopic verification of the presence of *Taenia* eggs, no amplification of the *Tso31* gene was found, suggesting that in these samples, the infecting species could be *T. saginata*. To rule out that the negative results were due to the presence of PCR inhibitors in the extracted DNA, considering that these were stool samples stored for more than 5 years in formalin-glycerol, the experiment was repeated with the addition of 20 ng of DNA of *T. solium* in 200 µl of the sample before performing total DNA extraction. In these spiked samples, it was possible to amplify the *Tso31* gene fragment, suggesting that the negative results were not due to PCR inhibitors (Fig. 3B). This test was performed on all negative samples (results not shown). y no se detectó ADN de *T. solium*

**Fig. 3.**
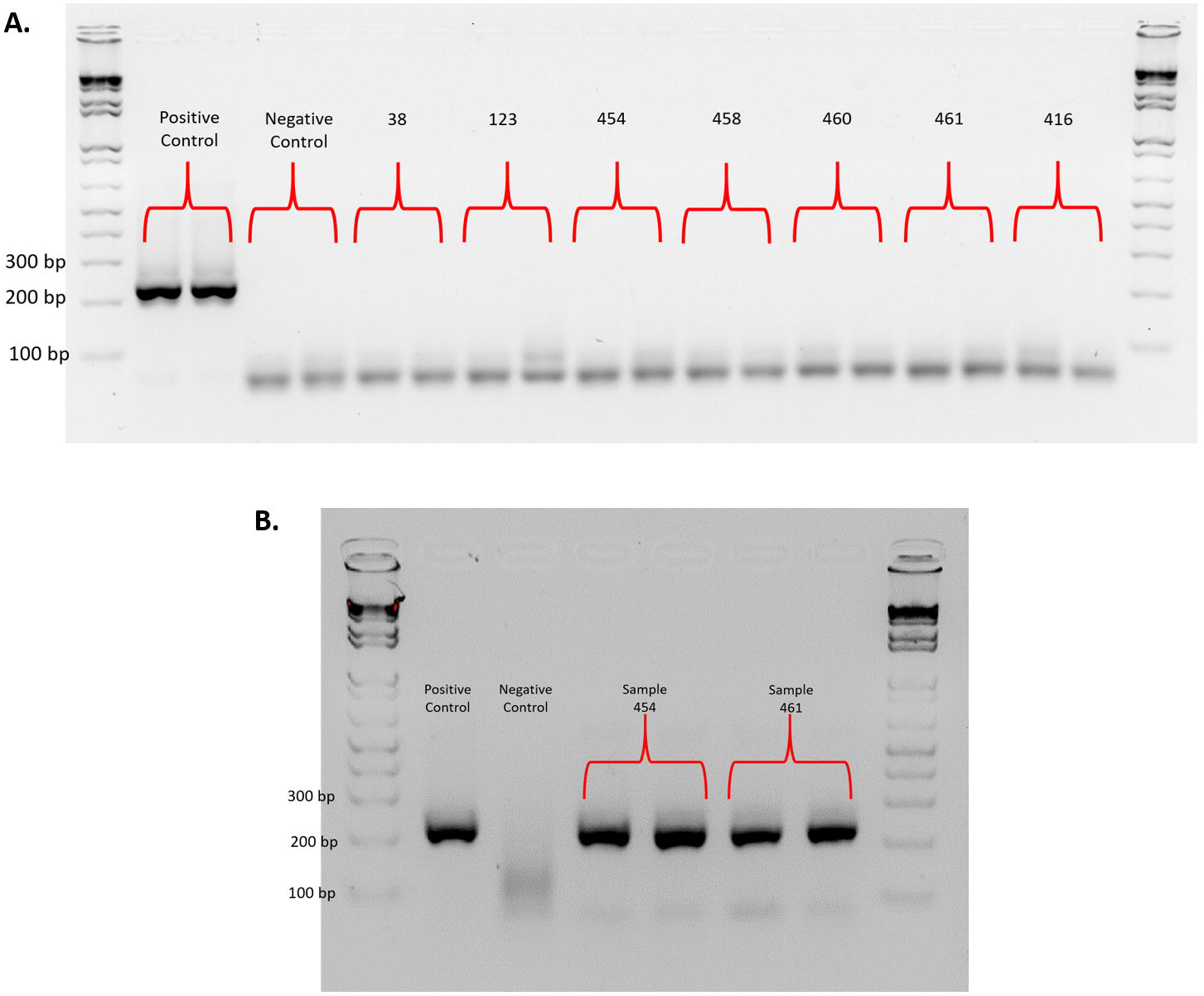
The PCR is negative in human fecal samples with parasitological verification of different intestinal parasites. **A**. The results of the nested PCR of the *Tso31* gene on DNA extracted from samples of human fecal material preserved in formalin-glycerol with parasitological verification by microscopy. <u>Sample 39</u>: *Taenia spp. Entamoeba coli* cysts; <u>Sample 123</u>: eggs of *Hymenolepis diminuta, E. coli* cysts, *Giardia* cysts, *Ascaris* eggs, *Trichuris trichiura*; <u>Sample 454</u>: *Uncinaria* cysts, *B. hominis, Giardia* cysts; <u>Sample 458</u>: *Ascaris* eggs, *Trichuris trichiura, E. coli* cysts, *E. hartmani* cyst, *Chilomastix mesnili*; <u>Sample 460</u>: *Hymenolepis nana, E. coli* cysts, *E. nana, Giardia* cysts, *E. histolytica* complex; <u>Sample 461</u>: *Ascaris* eggs; <u>Sample 416</u>: *Uncinaria* eggs, *Ascaris* eggs, *Trichuris trichiura, E. coli* cysts, *Giardia* cysts. All experiments were performed in duplicate. Positive control: DNA extracted from *T. solium* in adult form recovered from hamster. Negative control: reaction mixture without template. **B**. Evaluation of the presence of PCR inhibitors in the DNA extracted from human fecal material samples and preserved in formaldehyde-glycerol; 20 ng of *T. solium* DNA in 200 µl of sample before performing total DNA extraction.

## DISCUSSION

Nested PCR of a fragment of the *Tso31* gene can specifically detect DNA from *T. solium* in stool samples without showing cross-reaction with other species of the genus *Taenia* or other intestinal parasites of public health importance in Colombia. Therefore, this is an alternative for the detection of the parasite that can be considered in laboratories that have the necessary infrastructure to perform molecular biology tests.

The PCR detection limit we found is similar to that reported in a study that amplified fragments of the *Cox1, Nad5*, and 12S genes in a general PCR protocol for the detection of DNA of parasites of the class *Cestoda*. The authors found that the PCR detection limit could vary by several orders of magnitude, depending on the type of sample and the molecular target chosen, similar to what was found in the present study (12).

No PCR cross-reaction was found with DNA from other intestinal parasites or with DNA extracted from adult forms of *T. saginata* and *T. crassiceps*. The results showed the amplification of a single 234-bp fragment only in the samples with *T. solium* DNA. However, Vargas-Calla et al. (2019) evaluated whether the *Tso31* gene was conserved in other *Taenia* species and found that PCR of it was negative in DNA from *T. omisa, T. hydatigena, T. pisiformis*, and *T. taeniaeformis* but was positive in *T. multiceps*; the authors suggested that the *Tso31* gene sequence is not exclusive to *T. solium* (13). Considering the above, it is important to continue evaluating the possible cross-reactions of this test with other, related parasites.

For the PCR technique, 100% specificity and 97-100% sensitivity are reported (8), whereas the sensitivity of the microscopic method can vary between 3.9% (5) and 52.5% (6). In addition to the high sensitivity and specificity, the PCR method offers the advantage of differentiating between *T. saginata* and *T. solium*, which favors the identification of the infecting species, as in the case of the samples included in the present study, which, despite microscopic verification of the presence of *Taenia* eggs, were negative for *T. solium*.

Immunodiagnosis based on the detection of *T. solium* antigens in stool samples is another method that has been suggested to overcome the limitations of microscopic diagnosis. For this method, sensitivity rates between 96.4% and 98% and specificity rates between 99.2% and 100% are reported (5, 14). However, the level of species-specific specificity reported for these tests is only reached after the recovery of secretion/excretion antigens obtained by the infection of animal models with larvae of different *Taenia* species and the recovery of the parasite in its adult stage (14). This can be avoided with the nested PCR evaluated in this study, but this technique can only be applied in laboratories equipped with the necessary infrastructure.

The possibility to bring parasite DNA-based detection tests, such as PCR, to the patient point of care can be achieved through the development of loop-mediated isothermal amplification techniques. For this type of test, a sensitivity of 88.4% is reported for the diagnosis of *T. solium* infections (15). However, this method requires further development to achieve better operational characteristics (16).

One of the challenges of this study was the extraction of DNA from various types of samples, especially those preserved in formalin. Formalin-fixed biological samples stored in biological banks are of great importance for research based on genomic resources. However, the quality of the DNA is very poor due to the formalin-induced fragmentation and cross-linking of DNA with proteins (10). It is difficult to predict the quality of the extracted DNA because the levels of degradation and cross-linking with proteins may depend on factors such as pH, temperature, the ionic strength of the buffer, and the storage time of the samples under these conditions (11).

Nested PCR of the *Tso31* gene is a useful method for detecting *T. solium* DNA in stool samples and could be adopted by programs to control and/or eliminate the taeniasis/cysticercosis complex.

## Data Availability

Data will be available in the final publication and on request with the authors of the manuscript

## Acknowledgments

The authors thank María Teresa Herrera and Lyda Muñoz for their support in the laboratory of the Parasitology Group of the Public Health Research Directorate of the National Institute of Health of Colombia.

